# Childhood trauma as a mediator between autistic traits and depression: evidence from the ALSPAC birth cohort

**DOI:** 10.1101/2024.07.12.24310340

**Authors:** Jack F G Underwood, Paul Madley-Dowd, Christina Dardani, Laura Hull, Alex S F Kwong, Rebecca M Pearson, Jeremy Hall, Dheeraj Rai

**Affiliations:** Neuroscience and Mental Health Innovation Institute, Cardiff University, United Kingdom; Population Health Sciences, Bristol Medical School, University of Bristol, United Kingdom; MRC Integrative Epidemiology Unit (IEU), Bristol, United Kingdom; National Institute of Health Research Biomedical Research Centre, University of Bristol, United Kingdom; Division of Psychiatry, Centre for Clinical Brain Sciences, University of Edinburgh, United Kingdom; Department of Psychology, Manchester Metropolitan University, Manchester, United Kingdom; Avon and Wiltshire Partnership NHS Mental Health Trust, Bristol, United Kingdom

**Keywords:** ALSPAC, autism, depression, childhood trauma

## Abstract

**Background:** Autism and autistic traits have been associated with greater risk of childhood trauma and adulthood psychopathology. However, the role that childhood trauma plays in the association between autism, autistic traits and depression in adulthood is poorly understood.

**Methods:** We used a UK-based birth cohort with phenotype and genotype data on autism, autistic traits, childhood trauma and depression in up to 9,659 individuals prospectively followed up from birth until age 28 years. Using mixed-effects growth-curve models, we assessed trajectories of depression symptoms over time according to the presence or absence of autism/ autistic traits and explored whether these differed by trauma exposure. We further investigated the association between autism/ autistic traits and depression in adulthood using confounder-adjusted logistic regression models and undertook mediation analyses to investigate the relationship with childhood trauma.

**Results:** All autism variables demonstrated increased depressive symptom trajectories between ages 10-28 years. Social communication difficulties (SCDs) were the most strongly associated with a depression diagnosis in adulthood (age 24 OR= 2.15; 95%CIs: 1.22-3.76). Trauma and autistic traits combined to further increase depression symptom scores. Mediation analyses provided evidence for direct pathways between autistic traits and increased risk of depression alongside indirect pathways through increased risk of trauma.

**Conclusions:** Autism/ autistic traits increase the odds of experiencing childhood trauma and of being diagnosed with depression at age 18 and 24. Depressive symptom trajectories emergent in childhood persist into adulthood. The combined effect of SCDs and childhood trauma is greater than the individual exposures, suggesting worse depression symptomatology following trauma in individuals with SCDs.

**Lay abstract:** Autism and autistic traits are linked to higher chances of experiencing traumatic events in childhood and having mental health problems as an adult. However, we don’t understand how childhood trauma influences the link between autistic traits and depression in adulthood.

Using data from a large birth cohort study in the UK, we tracked up to 9,659 people from birth until age 28 years. We looked at the links between autism, autistic traits, childhood trauma, and depression.

We found that all features related to autism were associated with more symptoms of depression between the ages of 10 and 28. Among these, difficulties in social communication were the most strongly connected to depression when they reached adulthood, more than doubling the risk.

We discovered that when childhood difficulties and autistic traits came together, the risk of experiencing depression symptoms increased even more. We showed that autistic traits raised the chances of depression both directly and indirectly, by increasing the chance of experiencing childhood trauma.

Taken together, our research shows that autism and autistic traits increase the likelihood of experiencing childhood trauma and being diagnosed with depression at ages 18 and 24. Signs of depression that appear in childhood tend to persist into adulthood. The combination of social communication difficulties and childhood trauma has a stronger impact on depression symptoms, indicating that people with these traits may experience more severe depression after facing traumatic events.

## Background

Autism spectrum disorder (henceforth autism, using identity-first language, per community preference ^1–3)^ is a lifelong neurodevelopmental condition characterised by difficulties with reciprocal social communication and repetitive or restricted interests and behaviours ^4^. Its component traits extend into the general population ^5,6^, and potentially have distinct aetiologies conferred through environmental, common and rare genetic risk factors ^6–8^. Substantial evidence has suggested that depression occurs more commonly in autistic individuals, with pooled lifetime prevalence estimated at 11-37% ^9,10^, and some evidence of shared genetic architecture between autism and depression (*r*_g_ =0.412; *p*=1.40 x10^-25^) ^11,12^. In these individuals severity of social communication difficulties has been identified to be a core link to depression symptom severity and a key driver of quality of life ^13,14^.

Extensive work has suggested a robust association between childhood trauma and mental health difficulties in later life ^15–19^. This is particularly strong for depression, with odds ratios for later depressive illness following any childhood trauma ranging from 1.4 to 2.9 in the general population ^20–23^. Linkage studies of educational and healthcare records have shown that autistic children are at an elevated risk of maltreatment [odds ratio 1.86 (95%CI 1.36-2.52; p<0.001) in autistic children compared to matched general population controls], reinforcing results of qualitative and quantitative work in health care settings ^24–28^. It seems therefore that autistic individuals are at an elevated risk of childhood trauma and subsequent depression, however studies are limited. In earlier work we reported that bullying explained a substantial proportion of the increased risk of depression in autism up to age 18 ^14^. A subsequent survey of 902 parents of autistic children suggested lower socioeconomic status and poorer parental mental health were linked to elevated numbers of childhood trauma, and that childhood trauma was linked to later wellbeing including an association with increased anxiety and depression ^29^. This supports studies which identified that bullying is associated with ADHD and depression in autistic children ^30^. Genetically informed designs in large adult samples (UK Biobank), suggest that self-harm and suicidal behaviour and ideation have genetic correlations with autism, and this effect may be moderated by childhood trauma ^31^. Here we substantially expand previous work, using an additional 10 years of follow-up data in the ALSPAC cohort to assess the association between autism and depression up to 28 years of age, account for potential genetic confounding, and explore whether any associations are mediated by childhood traumas.

## Methods

### Study design and participants

We used data from the ALSPAC birth cohort, which contains detailed prospectively-collected information on parents and children from pregnancy through to adulthood ^32–34^. ALSPAC recruited pregnant women from Bristol and surroundings (Avon), UK, with expected dates of delivery 1st April 1991 to 31st December 1992, with an initial number of pregnancies enrolled of 14,541 (Figure 1).

Further recruitment and enrolment increased data collected after the age of seven to 15,447 pregnancies, with 14,901 children alive at one year of age. 26 participants withdrew consent and are not included in this study. We included one child per multiple birth pregnancy to ensure statistical independence of observations (186 removed). Of the remaining children, eligibility criteria for inclusion in the current study included having at least one measure of autism diagnosis or autism-associated trait and at least one measure of depression or depressive symptoms at any time point (Figure 1).

Study data were collected and managed using REDCap electronic data capture tools hosted at the University of Bristol. REDCap (Research Electronic Data Capture) is a secure, web-based software platform designed to support data capture for research studies ^35^. Further information on the ALSPAC cohort is available on the study website (http://www.bristol.ac.uk/alspac)^32,33^, which contains details of all the data that is available through a fully searchable data dictionary and variable search tool (http://www.bristol.ac.uk/alspac/researchers/our-data/).

### Ethical approval

Ethical approval for the study (project B4018) was obtained from the ALSPAC Ethics and Law Committee and the Local Research Ethics Committees. Informed consent for the use of data collected via questionnaires and clinics was obtained from participants following the recommendations of the ALSPAC Ethics and Law Committee.

**Figure 1:**
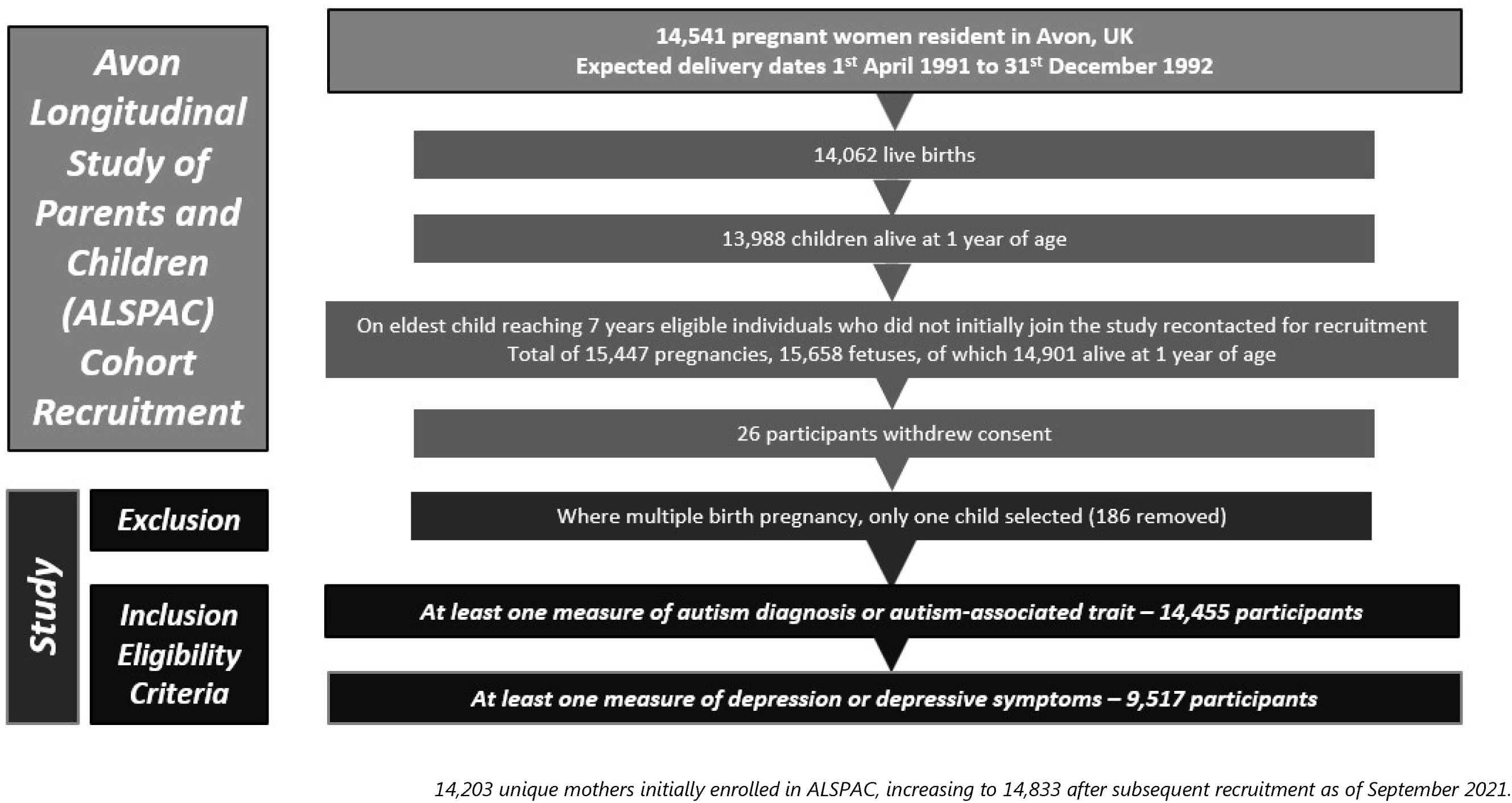
Flow chart of recruitment process to ALSPAC and the present study.

### Measures

We briefly describe information on our exposure, outcome, mediator and confounder variables here. Further details of all measures can be found in the Supplementary Material.

### Ascertainment of autism and autism-associated traits (exposure)

We utilised measures for autism diagnoses, autism associated traits and a polygenic score (PGS) for autism. Autistic children have previously been identified in the ALSPAC cohort using a multi-source approach, including review of clinical and educational records against International Classification of Diseases, 10th Revision [ICD-10] criteria ^14,35,36^. We included a measure of broad autism phenotype, the autism factor mean score, derived using factor analysis in ALSPAC as the mean of seven factors predictive of an autism diagnosis ^37^. We further used four independent trait predictors of autism which have been identified as being the best of 93 measures to predict ICD-10 autism diagnosis ^37^: the social communication difficulties trait (SCDs) - assessed at age 7 years via the Social Communication Disorder Checklist (SCDC) ^38^; the speech coherence trait (SC) - assessed at age 9 years via a subscale of the Children’s Communication Checklist ^39^; the repetitive behaviour trait (RRB) - assessed at age 5 years via parental questionnaire response ^40^; and the low sociability trait (LS) - assessed at age 3 years via a subscale of the Emotionality, Activity and Sociability Temperament Scale ^41^. For each autism measure the top decile scores were used to indicated case status in a binary measure of the trait. Finally, for the 6,380 children with genome-wide association (GWAS) data, we generated polygenic scores (PGS) for autism using standard QC methodologies at 13 p-thresholds in PLINK following the approach described in Ripke *et al* ^42^, using the 2019 Grove *et al* Psychiatric Genomics Consortia for Autism Spectrum Disorder dataset as a discovery sample ^12^. A threshold *p*-value of 0.5 was used for analyses as it explained maximum liability in our dataset ^42–45^.

### Ascertainment of depression (outcome)

We investigated both depressive symptom scores and formal clinical diagnosis of depression. The Short Mood and Feelings Questionnaire (SMFQ) ^46^, designed to measure depressive symptoms in children and adolescents, was administered at 11 time points between ages 10-28 years via postal questionnaires or in clinics ^47^. The computerized version of the Clinical Interview Schedule–Revised (CIS-R) ^48^ was also administered at ages 18 and 24 years to identify individuals with an ICD-10 diagnosis of depression.

### Ascertainment of childhood trauma (mediator)

The childhood trauma measures used in this study have been described in detail elsewhere ^49^. In brief, we used a contemporaneous measure between ages 11-17 based on responses to 57 questions from mothers and children in prospective questionnaires and interviews about a range of potential childhood traumas covering the following categories: domestic violence, physical abuse, emotional abuse, emotional neglect, sexual abuse and bullying. These measures were supplemented with retrospective self-reported questionnaire data obtained at age 22, pertaining to events between ages 11-17 ^49^.

### Potential confounders

We considered the following covariates as they are associated with both autism and depression: (1) child sex, (2) maternal parity, (3) maternal occupational class, (4) mother’s highest educational attainment, (5) financial problems when the child was 8 months old, (6) maternal age at delivery, (7) maternal Crown-Crisp anxiety score at 18 weeks’ gestation and 8 weeks after delivery ^50^, (8) maternal antenatal and postnatal depression symptoms scale ^51^, and (9) accommodation type ^14^. We further examined potential genetic confounding of associations between autism, depression and childhood trauma using a PGS for depression generated using the 2018 Psychiatric Genomics Consortia for Major Depressive Disorder dataset as the discovery sample following the same methodology as for the autism PGS ^52^.

### Statistical analysis

**Figure 2:**
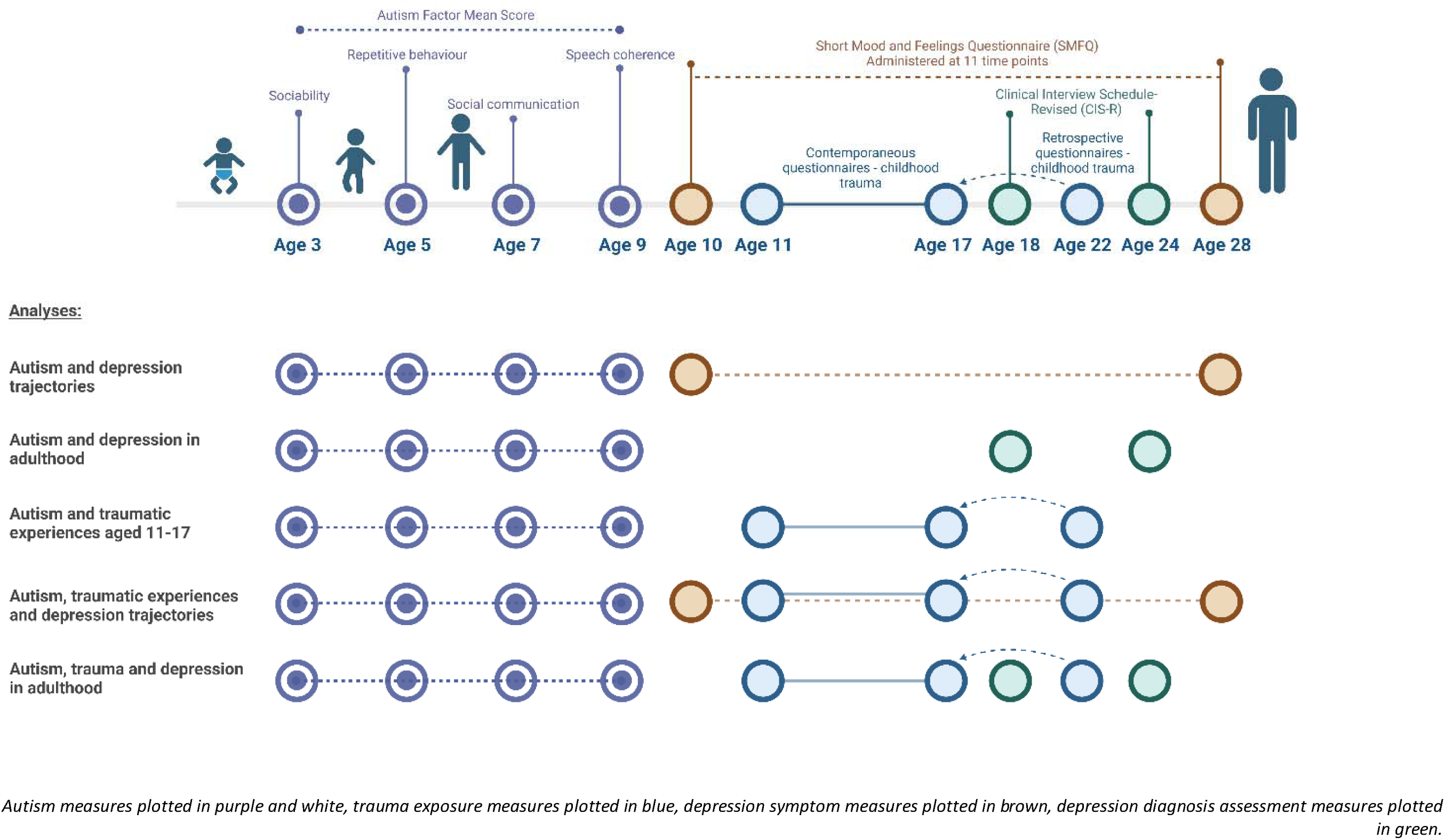
Plot of analyses undertaken by demonstrating timeline of measures and outcomes incorporated for each analysis.

We provide descriptive statistics of the sample separated by presence of an autism diagnosis or autism-associated trait. We briefly describe the statistical analysis below with further details of all models and derivation of trajectories provided in the Supplementary Material. A visual aid for all analyses is presented in Figure 2. All analyses were conducted using Stata/MP 17.0 (34).

We first examined trajectories of depressive symptoms (continuous SMFQ scores) between ages 10-28 years among those with and without each autistic trait, estimated using mixed-effects growth curve models. To explore the influence of childhood trauma on depression trajectories we repeated these models using four categories representing the combination of the presence or absence of both autistic traits and trauma variables ^53^.

We then assessed the association between autistic traits and i) ICD-10 depression diagnosis at age 18 and 24 and ii) each trauma type at ages 11-17 using logistic regression. We ran models unadjusted, adjusted for all potential confounders and again with further adjustment for depression PGS in those with genetic data. Finally, we assessed whether associations between autistic traits and depression diagnosis were mediated by trauma. Models were fit for all autistic traits with an identified association with depression diagnosis, and all trauma types associated with the autistic trait. We used the parametric g-formula and Monte Carlo simulations to estimate the natural direct effect (NDE) of autistic traits on depression, and the natural indirect effect (NIE) that was mediated via trauma, using the g-formula package in STATA ^54^.

We provide details of our missing data assessment in the Supplementary Methods. To account for potential bias from missing data in logistic regression, we performed multiple imputation with chained equations using 100 imputations and 25 burn-in iterations, with both imputed and complete case results presented ^55–57^.

## Results

The total sample size was 9,517 (47.5% male). Full characteristics of our study sample, including auxiliary and outcome variables are listed in Supplementary Table S6. Those with an autism diagnosis or in the top decile for autism trait groups were more likely to be male and had a consistently higher prevalence of reported childhood trauma for all trauma domains, particularly bullying and victimisation, excluding sexual abuse.

When assessing trajectories of depression symptoms, participants with an autism diagnosis or elevated autism trait measures in childhood had higher model-predicted scores (indicating greater depression) at age 10 (Figure 3, Supplementary Table S7 and Supplementary Figure S1). These elevated symptom scores continued across all time points, demonstrating a consistent uplift in trajectory through to adulthood, although with large overlaps of confidence intervals at many ages. The greatest increase in model-predicted depression symptoms across childhood and into adulthood was seen in those scoring highest for SCD (Supplementary Table S7), followed by those with the highest autism factor mean score.

**Figure 3:**
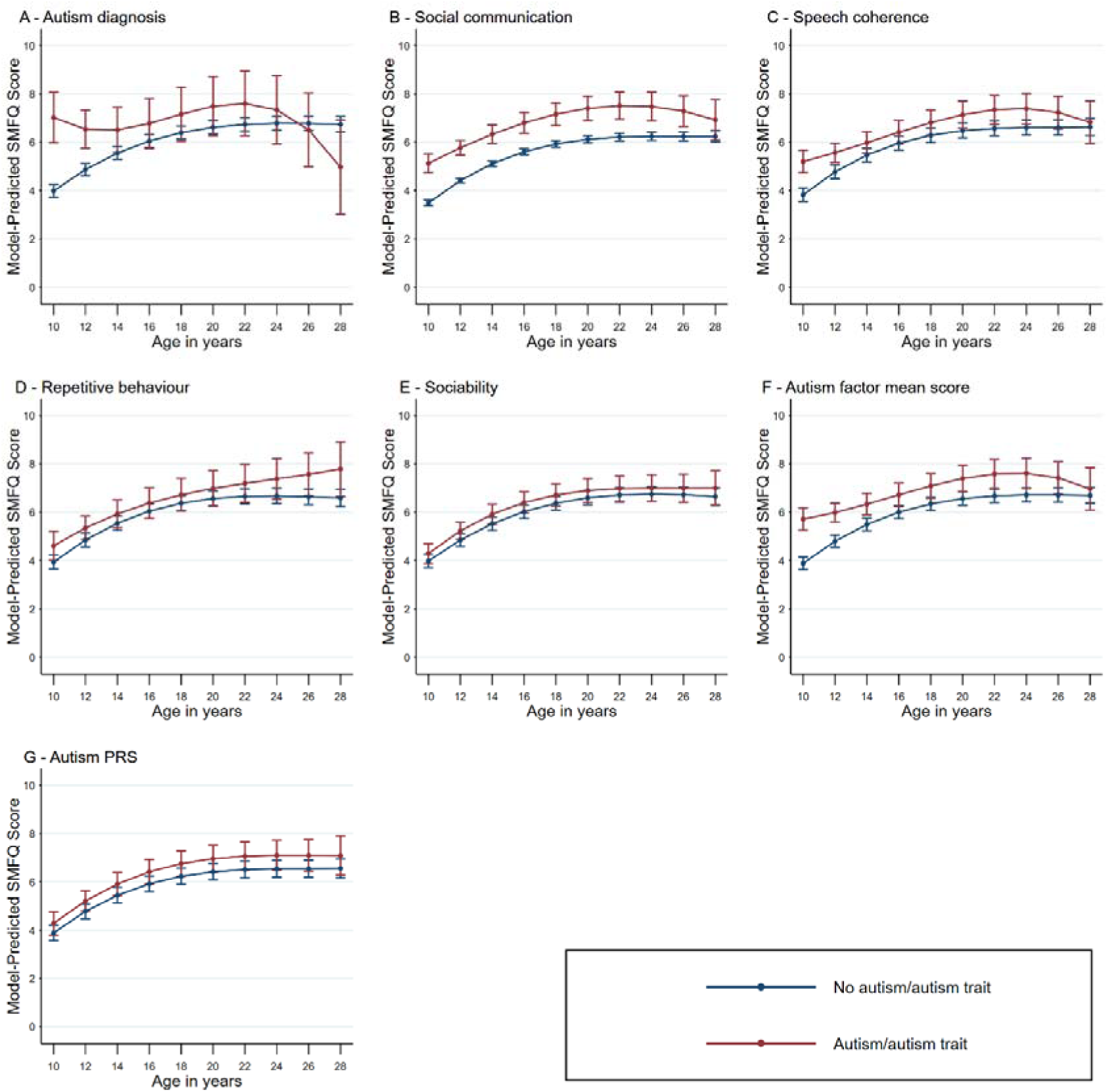
Trajectories of depressive symptoms between age 10 and 28 according to the presence or absence of each autistic trait measure.

Both SCDs and exposure to any trauma resulted in higher model-predicted depression symptoms scores across all age ranges (10-28) compared to those with no trauma or lower SCDs (Figure 4, Supplementary Table S8). When individuals with SCDs had experienced trauma, their scores were generally predicted to be greater than any other individual group, although trajectories varied and had large confidence intervals. This pattern applied across all individual trauma sub-domains and autism trait variables, with notable increases in depression scores between ages 12-20 of those with social communications difficulties who had experienced sexual or emotional abuse (Supplementary Results Figures S2-7).

**Figure 4:**
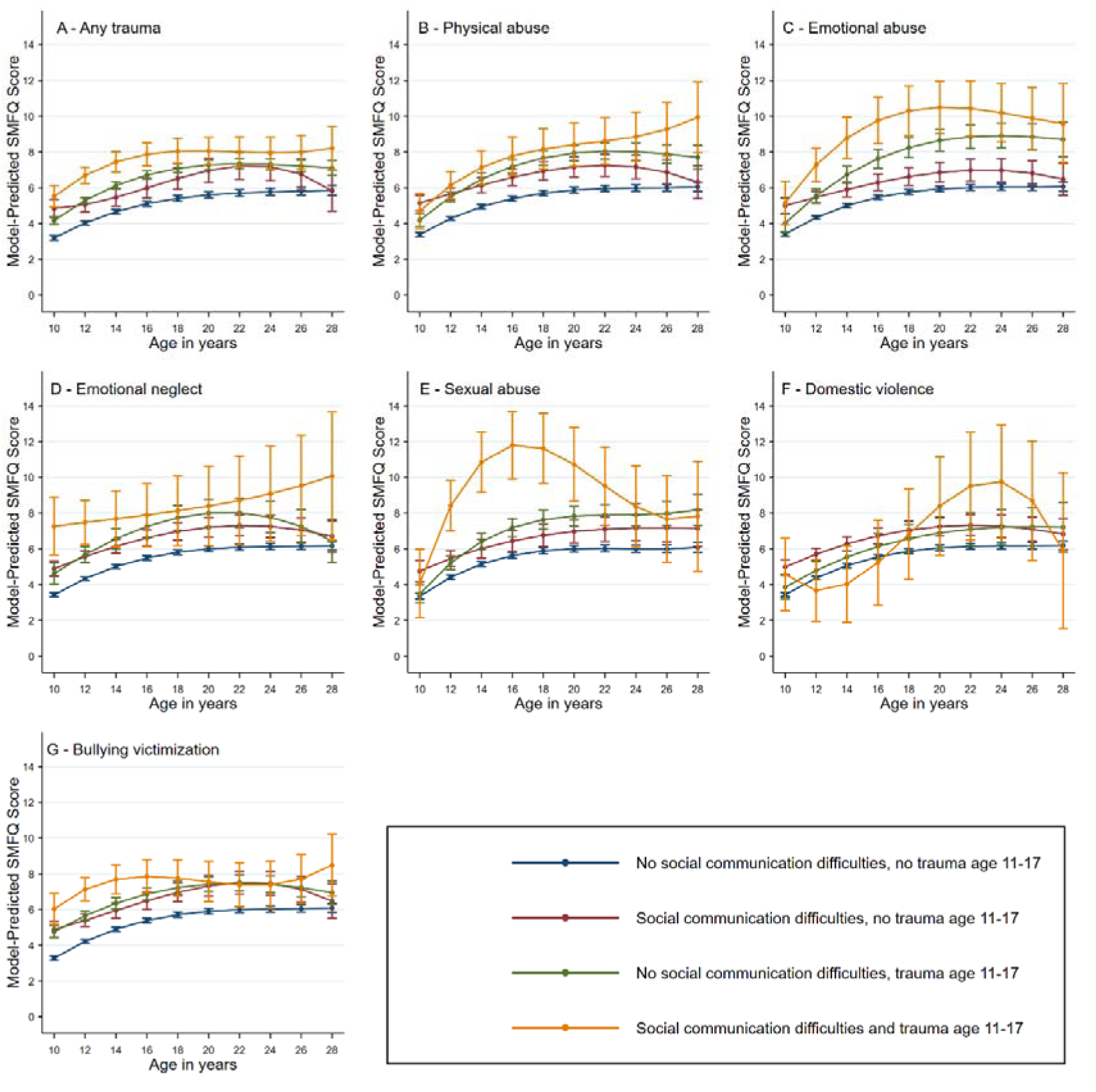
Trajectories of depressive symptoms between age 10 and 28 according to the presence or absence of the social communication trait and each trauma measure.

In logistic regression models of depression diagnosis, individuals with SCDs had greater odds of depression at age 18 (imputed sample; adjusted odds ratio [adj.OR]=1.76, 95%CI 1.23-2.53; Table 1) and 24 years of age (imputed sample; adj.OR=1.60, 95%CI 1.07-2.39) than those without SCDs.

**Table 1:** Odds ratios for depression diagnoses at age 18 and 24 ascertained using the CIS-R by autism/autistic traits.

| Exposure | Model | Complete records analysis |  |  |  |  |  | Multiple imputation analysis |  |
| --- | --- | --- | --- | --- | --- | --- | --- | --- | --- |
|  |  | Age 18 |  |  | Age 24 |  |  | Age 18 | Age 24 |
|  |  | N | Cases | OR (95% CI) | N | Cases | OR (95% CI) | OR (95% CI) | OR (95% CI) |
| Autism | Unadjusted | 2943 | 216 | 0.37 (0.05-2.70) | 2524 | 252 | 0.67 (0.16-2.81) | 0.48 (0.13-1.78) | 1.31 (0.54-3.18) |
|  | Adjusted for confounders | 2943 | 216 | 0.45 (0.06-3.40) | 2524 | 252 | 0.89 (0.21-3.85) | 0.61 (0.16-2.33) | 1.61 (0.63-4.07) |
|  | Adjusted for confounders and depression PGS | 2262 | 161 | 0.63 (0.08-4.82) | 1909 | 181 | 1.13 (0.26-4.96) | 0.57 (0.15-2.19) | 1.52 (0.60-3.89) |
| Social communication | Unadjusted | 2633 | 190 | 1.56 (0.97-2.52) | 2241 | 207 | 1.79 (1.13-2.83) | 1.69 (1.21-2.37) | 1.59 (1.08-2.35) |
|  | Adjusted for confounders | 2633 | 190 | 1.71 (1.04-2.83) | 2241 | 207 | 1.86 (1.15-3.01) | 1.76 (1.23-2.53) | 1.60 (1.07-2.39) |
|  | Adjusted for confounders and depression PGS | 2054 | 144 | 1.82 (1.03-3.22) | 1734 | 153 | 2.15 (1.22-3.76) | 1.71 (1.19-2.46) | 1.56 (1.04-2.34) |
| Speech coherence | Unadjusted | 2651 | 189 | 0.61 (0.32-1.18) | 2247 | 207 | 1.48 (0.92-2.37) | 0.86 (0.56-1.32) | 1.36 (0.93-2.00) |
|  | Adjusted for confounders | 2651 | 189 | 0.64 (0.33-1.24) | 2247 | 207 | 1.62 (0.99-2.64) | 0.87 (0.56-1.37) | 1.38 (0.93-2.05) |
|  | Adjusted for confounders and depression PGS | 2061 | 143 | 0.61 (0.29-1.30) | 1730 | 155 | 1.86 (1.06-3.26) | 0.86 (0.55-1.35) | 1.37 (0.92-2.04) |
| Repetitive behaviour | Unadjusted | 2528 | 183 | 1.20 (0.62-2.33) | 2158 | 204 | 1.28 (0.67-2.45) | 1.24 (0.73-2.11) | 1.75 (1.09-2.81) |
|  | Adjusted for confounders | 2528 | 183 | 1.21 (0.61-2.42) | 2158 | 204 | 1.26 (0.65-2.46) | 1.24 (0.71-2.16) | 1.72 (1.04-2.83) |
|  | Adjusted for confounders and depression PGS | 1956 | 136 | 0.68 (0.24-1.94) | 1651 | 146 | 1.63 (0.74-3.58) | 1.22 (0.70-2.14) | 1.70 (1.03-2.80) |
| Sociability | Unadjusted | 2798 | 204 | 0.91 (0.56-1.47) | 2393 | 229 | 1.09 (0.71-1.68) | 0.84 (0.56-1.25) | 1.19 (0.86-1.64) |
|  | Adjusted for confounders | 2798 | 204 | 0.95 (0.58-1.53) | 2393 | 229 | 1.20 (0.78-1.87) | 0.86 (0.57-1.29) | 1.21 (0.87-1.69) |
|  | Adjusted for confounders and depression PGS | 2163 | 153 | 0.79 (0.42-1.46) | 1832 | 167 | 0.88 (0.49-1.58) | 0.85 (0.56-1.27) | 1.20 (0.87-1.67) |
| Autism factor mean score | Unadjusted | 2941 | 216 | 0.88 (0.50-1.55) | 2520 | 252 | 1.01 (0.60-1.69) | 1.06 (0.73-1.54) | 1.16 (0.79-1.70) |
|  | Adjusted for confounders | 2941 | 216 | 0.97 (0.54-1.74) | 2520 | 252 | 1.08 (0.63-1.86) | 1.13 (0.76-1.69) | 1.21 (0.81-1.80) |
|  | Adjusted for confounders and depression PGS | 2262 | 161 | 0.61 (0.28-1.37) | 1909 | 181 | 1.26 (0.66-2.43) | 1.14 (0.76-1.70) | 1.21 (0.81-1.81) |
| Autism PGS | Unadjusted | 2262 | 161 | 1.76 (1.14-2.72) | 1909 | 181 | 1.44 (0.93-2.25) | 1.70 (1.22-2.38) | 1.09 (0.77-1.53) |
|  | Adjusted for confounders | 2262 | 161 | 1.82 (1.16-2.85) | 1909 | 181 | 1.38 (0.88-2.16) | 1.70 (1.20-2.41) | 1.07 (0.75-1.51) |

<sup>a</sup> N for multiple imputation analysis = 9,517; N for PGS analysis = 6,380

Higher autism PGS was associated with increased risk of depression at age 18 but not at age 24. By contrast RRBs were associated with an increased risk of depression at 24 but not at age 18. None of the other autistic traits, autism diagnosis or autism factor mean score were clearly associated with a depression diagnosis at age 18 or 24, although 95%CIs were wide, probably reflecting small numbers.

Autism diagnosis plus a range of autistic traits were associated with increased risk of traumatic experiences between ages 11-17. SCDs were associated with increased odds of experiencing physical abuse (adj.OR=1.45, 95%CI 1.15-1.83; Supplementary Table S9), emotional abuse (adj.OR=1.99, 95%CI 1.50-2.63), and bullying (adj.OR=1.74, 95%CI 1.42-2.13). Elevated autism factor mean scores were associated with increased odds of experiencing emotional abuse (adj.OR=1.35, 95%CI 1.00-1.81), emotional neglect (adj.OR=1.92, 95%CI 1.41-2.61), and bullying (adj.OR=1.81, 95%CI 1.49-2.21), demonstrating the different patterns of trauma associated with each autism measure.

We further examined whether the associations between autistic traits in early childhood and depressive symptoms in adulthood were mediated by trauma in childhood (Supplementary Tables S10 & S11). We found evidence that trauma between ages 11 and 17 mediated associations between SCDs and depression diagnosis at age 18 (Natural Direct Effect (NDE): OR=1.54, 95%CI 0.96-2.49; Natural Indirect Effect (NIE): OR=1.09, 95%CI 1.03-1.13; Proportion Mediated (PM)=16.54%) and 24 (NDE: OR=1.67, 95%CI 1.02-2.71; NIE: OR=1.09, 95%CI 1.04-1.15; PM=14.84%).

## Discussion

In this longitudinal cohort study, we examined the interplay of autism, childhood trauma and depression symptom severity across development. We found that specific autistic traits, particularly SCDs in childhood, were associated with elevated depression symptom trajectories from age 10-28 and clinical diagnosis of depression in adulthood. Most autism variables analysed, including autism PGS but excluding low sociability, were associated with exposure to any childhood trauma. SCDs and autism factor mean score demonstrated the strongest association with any trauma and displayed specific autism trait to trauma subtype associations. These findings were robust to adjustment for potential confounders including polygenic scores for depression. Our trajectory analyses implied individuals with SCDs experience worse depression outcomes following trauma, as their depression symptoms scores were increased beyond either SCD or trauma independently.

The most common co-occurring conditions in autism are anxiety and depression, occurring at lifetime prevalence of 42% and 37% respectively ^10^. We previously demonstrated in the ALSPAC dataset that SCDs are a key driver of depression in autistic children and young people ^14^. This study extends on that work and is the first to show using a longitudinal methodology that depression symptoms associated with SCDs continue to be elevated into adulthood. These findings bolster a 2020 systematic review which postulated a model where SCDs confer depression risk in autistic individuals via social motivation, loneliness and isolation ^58^. Here we observed that elevated depression symptoms scores associated with some autistic traits fell from the age of 24 back to levels estimated in the wider sample. This likely reflects attrition from the study of those with the worst difficulties. An alternative hypothesis may be as autistic individuals grow into adulthood different features of their autism, such as RRBs, become relevant to risk of depression.

Considerable evidence suggests that autistic individuals experience both more traumatic experiences and experience them with greater intensity ^26,28,59,60^. We found differences in associations between autistic traits and trauma domains, with the strongest effect from SCDs, continuing into adulthood. SCDs may be especially important as individuals might consequently struggle understanding and communicating their emotions after trauma. This may make seeking support harder, with reports from experts by experience that neurodiverse reactions to trauma are often treated differently ^61^. The effect of trauma on subsequent mental health outcomes in autism has rarely been studied, despite evidence in the general population to suggest a dramatic impact ^20,27^. Studies predominantly investigate bullying in children ^14,29,30^, with a small number examining the full range of trauma domains or following up into adulthood ^31,60,62^. Two previous studies have attempted to model relationships between these exposures. Hollocks *et al.* used structural equation modelling in 115 participants from a population-based longitudinal study to demonstrate an association between childhood trauma and rates of emotional and behavioural symptoms in adulthood ^63^. Shin, Wright and Johnston concluded from longitudinal data in older adults (50+) that autism polygenic score alone could not predict co-occurring mental health outcomes, but was a factor in outcomes when combined with early life experiences ^64^. As here, autism PGS increased risk of mental health conditions when combined with childhood trauma ^65^. The finding of direct and indirect effects between trauma, autism and depression is therefore novel but unsurprising, and further investigation into how these mediate and moderate at different ages would be enlightening.

The current study improves on previous work through a longitudinal population-based cohort with a further 10 years of data into adulthood, and its incorporation of a broad range of confounders in modelling and multiple imputation to reduce the possibility of attrition bias. This population introduces limitations, as only a small number of participants (n=112) had an autism diagnosis, although this is consistent with the ∼1% population prevalence estimates of autism ^66^. We used further measures with prior evidence in ALSPAC to identify autistic traits and phenotypic features in a binarized manner across the whole population ^14,36^, however our estimates remain limited by the low prevalence of trauma in combination with these measures. To reduce exposure-mediator overlap we have ensured autistic traits are measured at prior time periods to trauma domains. The possibility of some reverse causation cannot be excluded; if participants unmeasured trauma exacerbated autistic traits or features (such as worsened social communication) and this aggravated depression symptomatology. There is also the possibility trauma itself causes the traits assessed, thus we are measuring the assessed phenotype consistent with an autism diagnosis. Although this is a longitudinal study the possibility of residual confounding cannot be excluded, and our estimates of the proportion mediated include considerable uncertainty.

## Conclusions

Our findings demonstrate that autism, autism genetic liability and most autistic traits were associated with increased depression symptoms from early childhood to age 28, increased odds of childhood trauma, and increased odds of clinical diagnosis of depression in early adulthood. Social communication difficulties are particularly important, conferring the greatest risk for childhood trauma and the greatest increase in depression symptoms. Exposure to childhood trauma further increases depression symptom scores in autistic individuals, with modelling demonstrating that the increased risk of depression observed in autism is partially indirectly caused by increased childhood trauma. These findings add to evidence demonstrating autistic people are at greater risk of childhood trauma, resulting in more severe depressive outcomes. Better understanding of trauma responses in autistic adults may guide targeted interventions, improving subsequent mental health.

## Data Availability

All scripts used to create the data and run analyses are available from https://github.com/pmadleydowd/Autism-childhoodtrauma-depression. ALSPAC data cannot be shared publicly due to ethical agreements. Data is available on application to ALSPAC through their website: http://www.bristol.ac.uk/alspac.

## Acknowledgements

We are extremely grateful to all the families who took part in this study, the midwives for their help in recruiting them, and the whole ALSPAC team, which includes interviewers, computer and laboratory technicians, clerical workers, research scientists, volunteers, managers, receptionists and nurses. We thank Dr Hein Heuvelman, for his earlier versions of statistical code updated for this work.

