## Supplementary Material for "Childhood trauma as a mediator between autistic traits and depression: evidence from the ALSPAC birth cohort"

### 1 Supplementary Methods

#### 1.1 Data Sharing Statement

All scripts used to create the data and run analyses are available from <https://github.com/pmadleydowd/Autism-childhoodtrauma-depression>. ALSPAC data cannot be shared publicly due to ethical agreements. Data is available on application to ALSPAC through their website: <http://www.bristol.ac.uk/alspac>.

#### 1.2 Ascertainment of autism diagnosis

The multi-source approach used to identify autism cases in ALSPAC comprised the following: review of clinical records of all children who had multidisciplinary assessment for a developmental disorder (validated against International Statistical Classification of Diseases, 10th Revision [ICD-10] criteria by a consultant paediatrician) (1); review of educational records of special education support provided for autism; and parental reports of an autism or Asperger syndrome diagnosis (2). The autism cases identified have been cross-validated against autism-associated trait measures (2,3), and the variable is associated with a genetic risk score for autism derived from an independent GWAS (Supplementary Figure 7) (4).

#### 1.3 Derivation of polygenic scores for autism and depression

We used PGSs for the outcome and not the exposure as any adjustment for an instrumental variable for the exposure can amplify bias from unmeasured confounding (5). We created an autism PGS variable using the 2019 PGC for Autism Spectrum Disorder GWAS as the discovery sample (6). We applied standard QC methodologies following the approach laid out in Ripke *et al.* including filtering on an expected allele frequency of >0.01, info score of >0.9 and excluding the MHC region (7). We then created a set of scores based on single-nucleotide polymorphisms (SNPs) that are associated with a depression diagnosis at 13 GWAS *p­-*value thresholds (.5 to 1e−7). Polygenic scores (PGS) for depression was calculated for genotyped ALSPAC children using summary data from the Wray *et al* 2018 Psychiatric Genomics Consortium (PGC) genome-wide association study (GWAS), with the same methodology as for the autism PGS (8). This GWAS sample was selected for it’s in-depth phenotyping and robust statistical methodology. The SNPs meeting the GWAS *p­-*value threshold of 0.5 maximally captured liability within our sample and were therefore used for analysis.

#### 1.4 Depression Measures

To investigate depressive symptoms in the ALSPAC dataset we used the Short Mood and Feelings Questionnaire (SMFQ) (9). This was designed to measure depressive symptoms in children and adolescents, and was administered at 11 time points between ages 10 and 28 years via postal questionnaires or in research clinics, specifically ages 10, 12, 13, 16, 17, 18, 21, 22, 23, 25 and 28, providing five additional time points to previous work (10,11). It has 13 items relating to low mood during the past 2 weeks, each with scores of 0 to 2. Individual item scores were summed, producing a 0 to 26 score range (11). To investigate clinical diagnosis of depression we used the Clinical Interview Schedule–Revised (CIS-R) (12). This is a fully structured psychiatric interview widely used in community samples including the UK Psychiatric morbidity surveys to estimate the national prevalence of depression and other common mental disorders (13). It was administered in ALSPAC in computerised form at age 18 and 24 and allowed us to identify individuals with an ICD-10 diagnosis of autism.

#### 1.5 Summary of childhood trauma variables

We used a contemporaneous measure of childhood trauma between ages 11 and 17 based on responses to 57 questions from questionnaires and interviews about a range of potential childhood traumas. These measures were supplemented with retrospective questionnaire data obtained at age 22, pertaining to events for that individual between ages 11 to 17 (14). Childhood trauma was derived from measures asking about domestic violence (presence of regular acts of physical violence taking place in the home), physical abuse (physical harm to the participant from caregivers or other adults), emotional abuse (emotional cruelty to the participant from caregivers or other adults), emotional neglect (caregivers not taking an interest in the participant’s life), sexual abuse (adults or older children forcing the participant into sexual activity, including attempts to do so), and bullying victimization (regular name-calling, blackmail, or assault by peers). Full information on trauma variables is available in Croft et al. 2019 (14). Contemporaneous measures recorded from participants and their caregivers between the ages of 5 and 11 were not used to reduce the overlap in measurement of autistic traits and exposure to trauma. Each type of childhood trauma was coded as present or not, and a single trauma variable was created representing exposure to any type of childhood trauma.

#### 1.6 Confounder definitions

We used the following confounders as covariates:

1. Child sex
2. Parity (≤1 child vs ≥2 children)
3. Maternal occupational class (manual vs nonmanual)
4. Mother’s highest educational attainment
5. Financial problems (occurrence vs non-occurrence of major financial problems)
6. Maternal age at delivery (in years)
7. Maternal Crown-Crisp anxiety score at 18 weeks’ gestation and 8 weeks after delivery (15)
8. Maternal antenatal (18 and 32 weeks’ gestation) and postnatal (8 weeks and 8 months) depression measured with the Edinburgh Postnatal Depression Scale (EPDS score ≥13) (16)
9. Accommodation type (detached house vs semidetached house vs flat).

#### 1.7 Statistical analytical methods

##### 1.7.1 Autism and depression trajectories

In calculating trajectories of depressive symptoms (continuous SMFQ scores) between ages 10-28 years among those with and without an autism diagnosis and for each autistic trait using mixed-effects growth curve models we accommodated individual differences in trends of depressive symptoms with age by including random intercept and random slope coefficients for age and age squared. We further included fixed effect linear, quadratic and cubic terms for age and their interaction with the autism measure to accommodate potential nonlinear trends, chosen as a compromise between complexity and parsimony, and learning from previous work by the authors in this dataset (17). We chose not to include random age coefficients at the cubic level as these effects can be quite small and result in convergence issues. These trajectories are therefore partially random as they allow everyone to have their own trajectory that varies from the population level trajectory, all the way to the quadratic level. These accommodations were also applied to mixed-effects growth models for the four-group analysis of presence or absence of autism or autistic traits and the presence or absence of childhood trauma (repeated for each trauma group). Data were not multiply imputed for trajectory modelling, and therefore we recognise that missing data may bias these results.

##### 1.7.2 Autism and traumatic experiences aged 11-17

We tested the association between autistic traits and trauma variables using logistic regression, unadjusted and adjusted for potential confounders. Models were repeated for each trauma type (domestic violence, physical abuse, emotional abuse, emotional neglect, sexual abuse and bullying) and autistic trait measure.

##### 1.7.3 Autism, trauma and depression in adulthood

We assessed whether any identified associations between autistic traits and depression diagnosis at age 18 and 24 years were mediated by the experience of any trauma, and each trauma type. Models were fitted for all autistic traits that were found to have an association with depression diagnosis, and all trauma types that were found to have an association with the autistic trait. Mediation analyses were performed using the g-formula package in STATA (18). We used the parametric g-formula and Monte Carlo simulations to estimate the natural direct effect (NDE) of autistic traits on depression, the natural indirect effect (NIE) that was mediated via trauma, and the proportion mediated. We performed models unadjusted and adjusted for all potential confounders. Corresponding 95% CIs were estimated using the standard errors from 1000 non-parametric bootstrap resamples.

##### 1.7.4 Multiple imputation

We decided a priori to perform multiple imputation (19,20) if bias due to missing data was deemed to be likely (i.e. associations between both outcome and exposure variables and complete records status are present in the data) using multiple imputation with chained equations (21) for logistic regression models only (19,22). All imputed datasets were created as part of the same procedure that included all outcomes, exposures, mediators and confounders in each prediction model. One hundred datasets were imputed using Stata’s *MI impute* command and estimates were combined across imputed datasets using Rubin’s rules implemented via Stata’s *MI estimate* command. We included auxiliary variables which were predictive of the unobserved missing values in the imputation model to make the missing at random assumption (23) more plausible, as required for unbiased estimation using multiple imputation. The repeated collection of SMFQ data over time in ALSPAC meant that data observed at one timepoint could be used as an auxiliary to predict missing values at other time points. Observed data on autistic traits and diagnosis were also used as auxiliary information for unobserved exposure information. Suitable auxiliaries were also selected for missing covariate and mediator variables from the ALSPAC resource as listed below. These auxiliary variables were included in all prediction models in order to make the missing at random assumption, required by standard multiple imputation implementation, more plausible. This assumption states that the probability of missing data is not dependent on unobserved information, conditional on the observed information. The auxiliary variables included parental marital status, weekly income, financial difficulties and use of a car during pregnancy for predicting missing socioeconomic variables. History of maternal depression and SMFQ scores acted as auxiliary variables for missing depression diagnoses, and each of the autism diagnoses and traits acted as auxiliary information for each other.

Auxiliaries utilised:

- History of maternal depression recorded at 12 weeks gestation – complete for 89.8% of the sample – binary measure
- Marital status – 8-42 week gestation – complete for 91.4% of the sample – categorical measure grouped as 1 "Never married", 2 "Previously married (currently unmarried)", 3 "1st marriage", 4 "2nd or 3rd marriage"
- Family weekly income – 33 months post pregnancy – complete for 72.8% of the sample – binary measure indicating weekly income of <£300 or ≥£300
- Financial difficulties in pregnancy – 32 weeks gestation – complete for 87.3% of the sample - Financial difficulties were measured using a self-report questionnaire during pregnancy. A score (ranging from 0 to 15) was derived from the sum of responses indicating the level of difficulty in affording food, clothing, heating, rent or mortgage and “things you will need for the baby”. Scores higher than 4 were indicated as having financial difficulties.
- Parental use of a car recorded during pregnancy - 8-42 week gestation – complete for 90.8% of the sample - binary measure

#### 1.8 Missing data assessment

##### 1.8.1 Calculation of missing data

We used the Treatment and Reporting of Missing data in Observational Studies framework (24) to make decisions about how to handle missing data. To assess whether bias in complete case analysis (where participants with missing data in any variable are excluded) was likely we compared the prevalence/means of exposure, outcome, mediator and confounder variables between those included in the sample and those excluded for missing data in any variable. We further performed logistic regression of being included in complete records analysis on each variable with adjustment for all potential confounders. Complete records analysis has been shown to be biased when the probability of missing data is jointly dependent on both the exposure and the outcome for logistic regression conditional on all adjusted variables (25). Below are provided descriptives and odds ratios for inclusion in complete records analysis (CRA) for analyses using the following exposures: autism diagnosis, social communication and autism PGS. We further provide plots of observed vs imputed values. We provide an FMI, a parameter-specific measure that is able to quantify the loss of information due to missing data, while accounting for the amount of information retained by other variables in a dataset (19,26). Values of FMI range between 0 and 1 with values close to 1 indicating high variability between imputed data sets meaning that the observed data in the imputation model does not provide much information about the missing values.

##### 1.8.2 Supplementary Methods Figure 1: Proportion exposed/with depression among the observed and imputed datasets

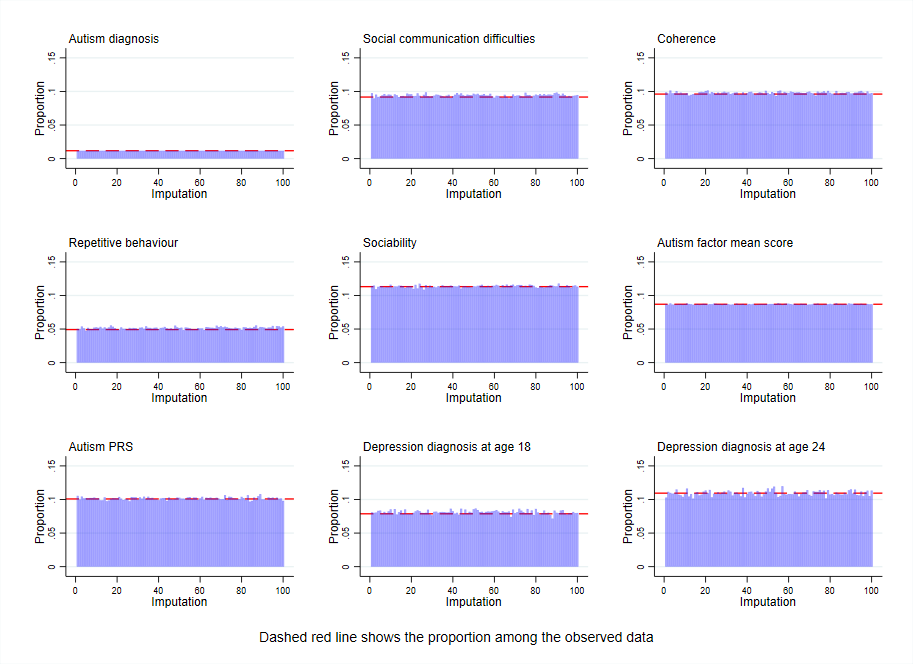

### 2 Supplementary Results Figures

#### 2.1 Supplementary Results Figure S1: Change in trajectories of depression symptom score between age 10 and 28 according to the presence or absence of each autistic trait

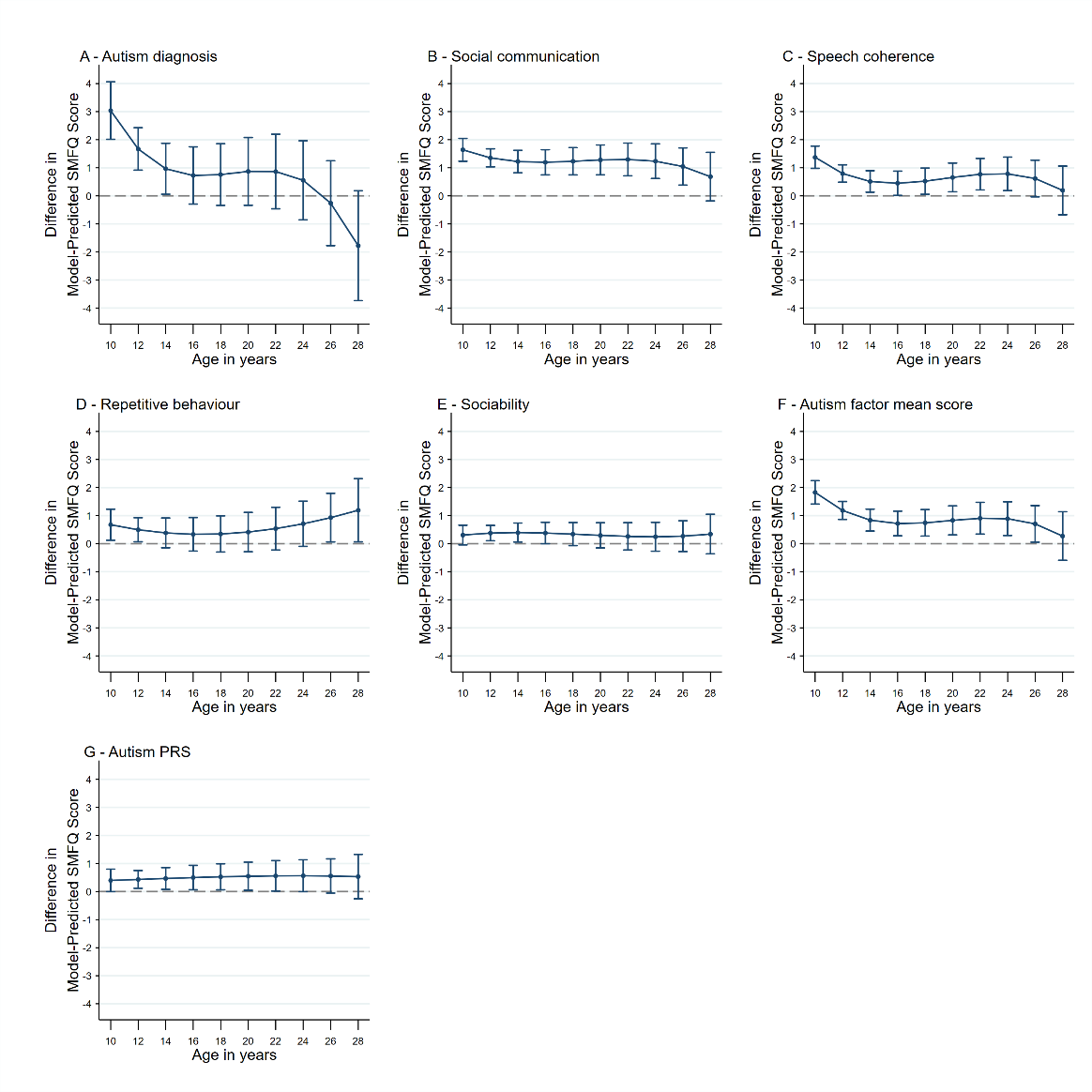

#### 2.2 Supplementary Results Figure S2: Trajectories of depressive symptoms between age 10 and 28 according to the presence or absence of an autism diagnosis and each trauma measure.

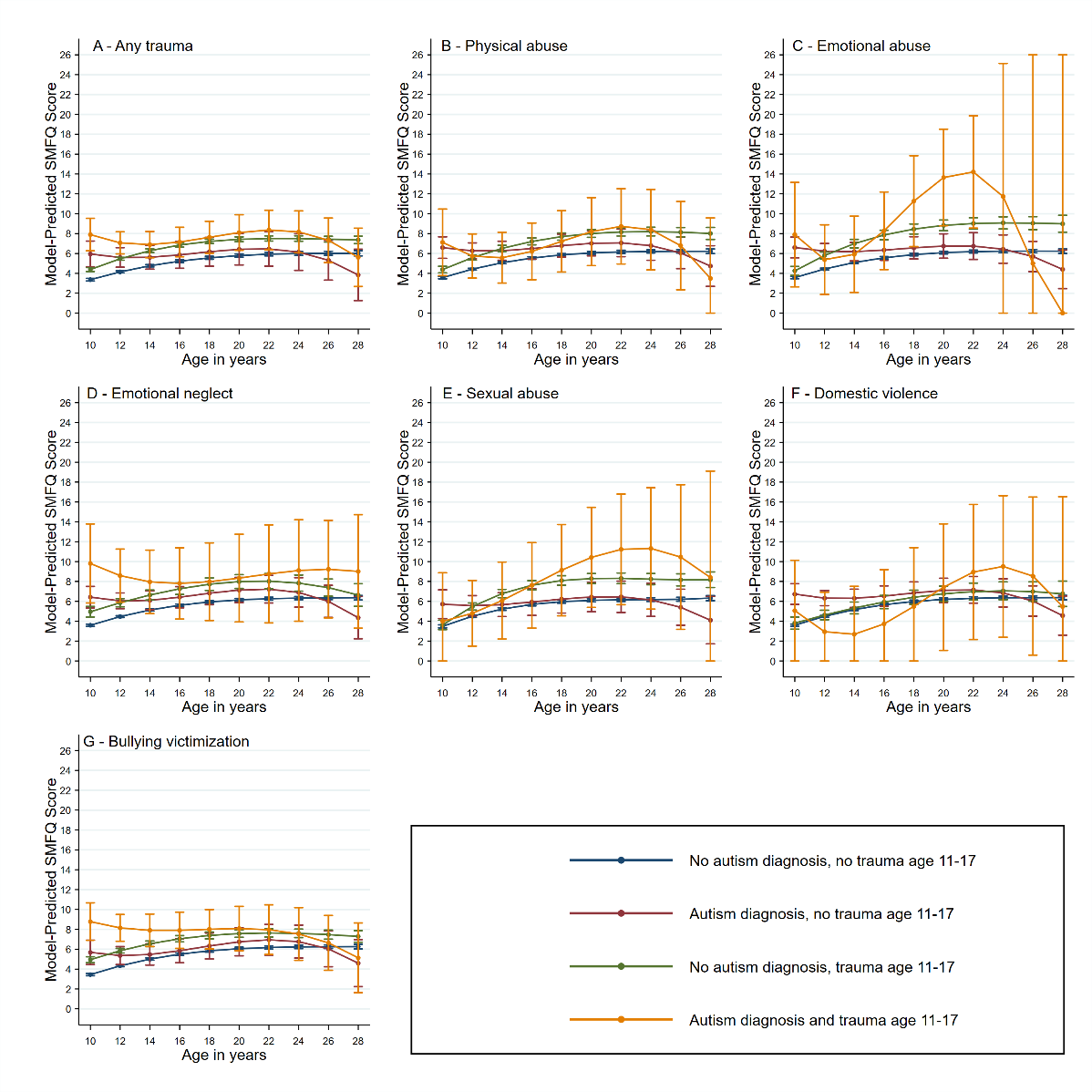

#### 2.3 Supplementary Results Figure S3: Trajectories of depressive symptoms between age 10 and 28 according to being in the highest decile of the autism factor mean score and presence or absence of each trauma measure.

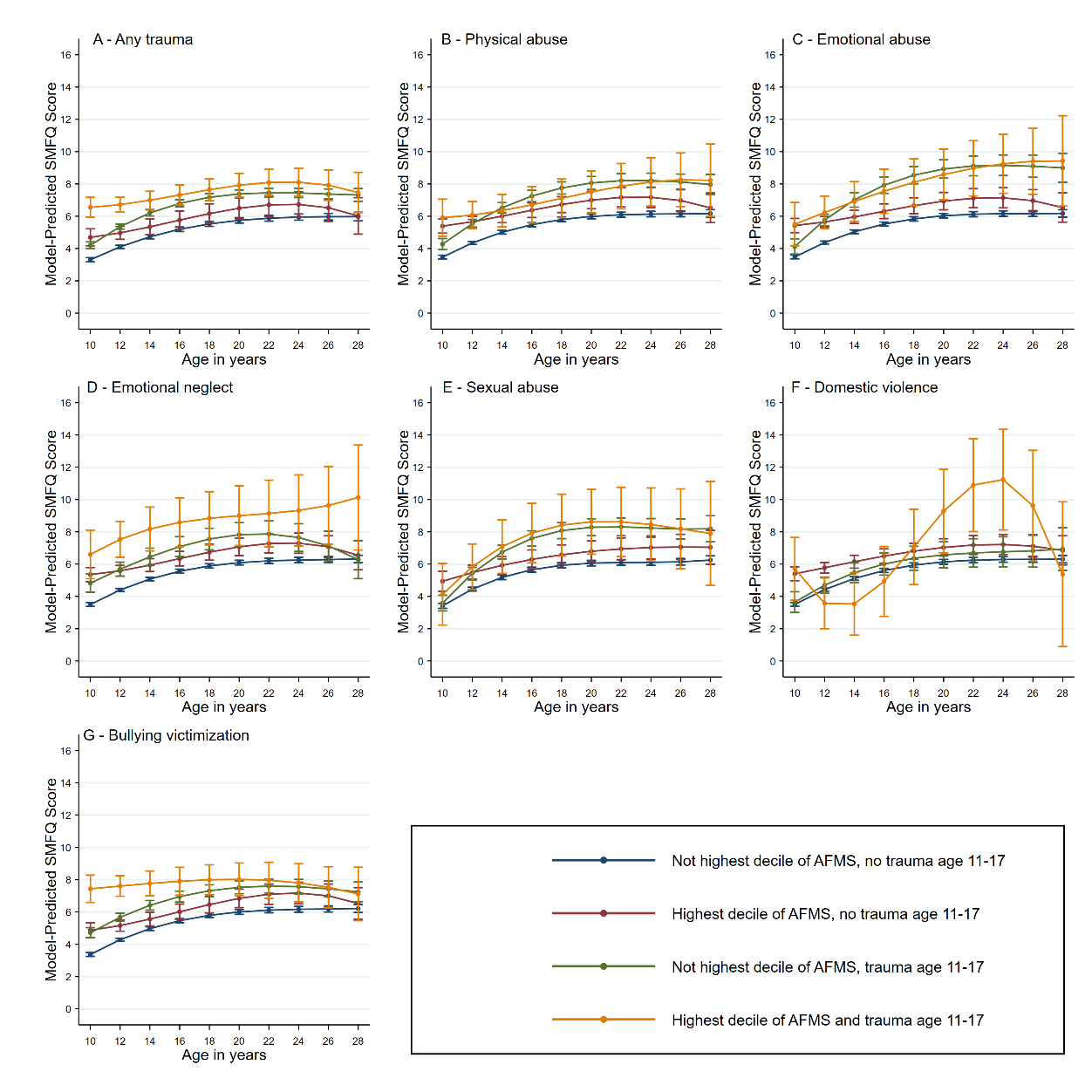

#### 2.4 Supplementary Results Figure S4: Trajectories of depressive symptoms between age 10 and 28 according to the presence or absence of the speech coherence trait and each trauma measure.

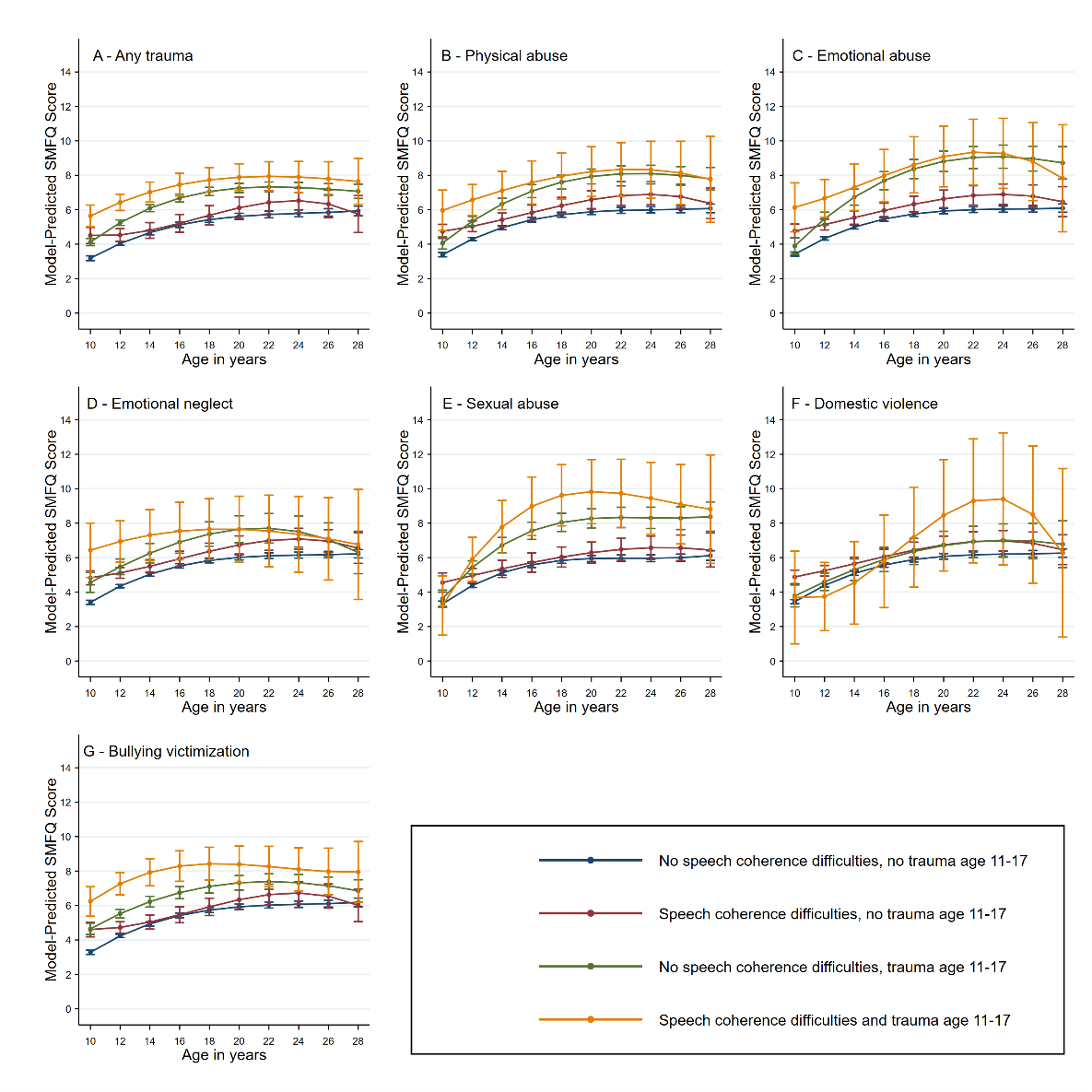

#### 2.5 Supplementary Results Figure S5: Trajectories of depressive symptoms between age 10 and 28 according to the presence or absence of the repetitive behaviour trait and each trauma measure.

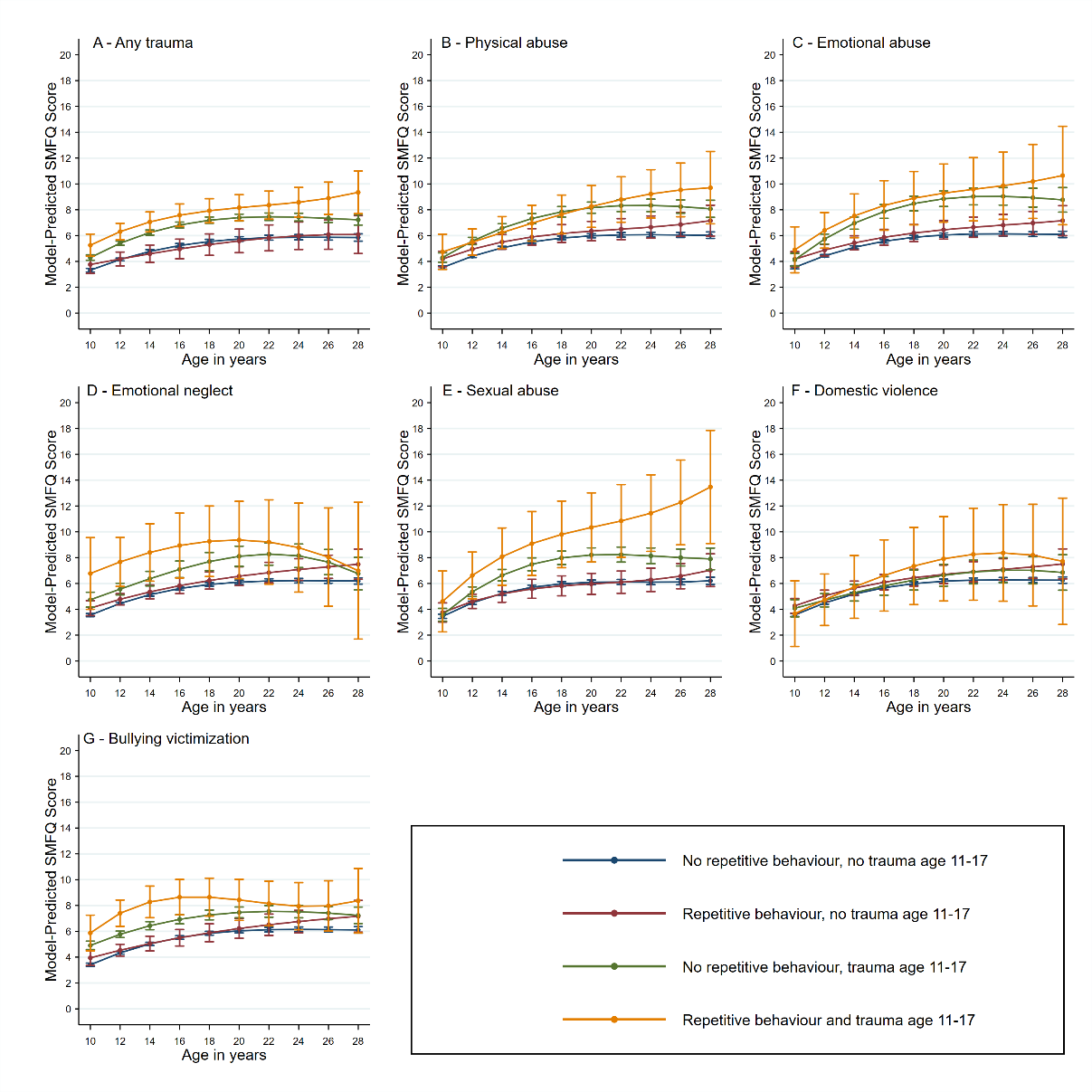

#### 2.6 Supplementary Results Figure S6: Trajectories of depressive symptoms between age 10 and 28 according to the presence or absence of the low sociability trait and each trauma measure.

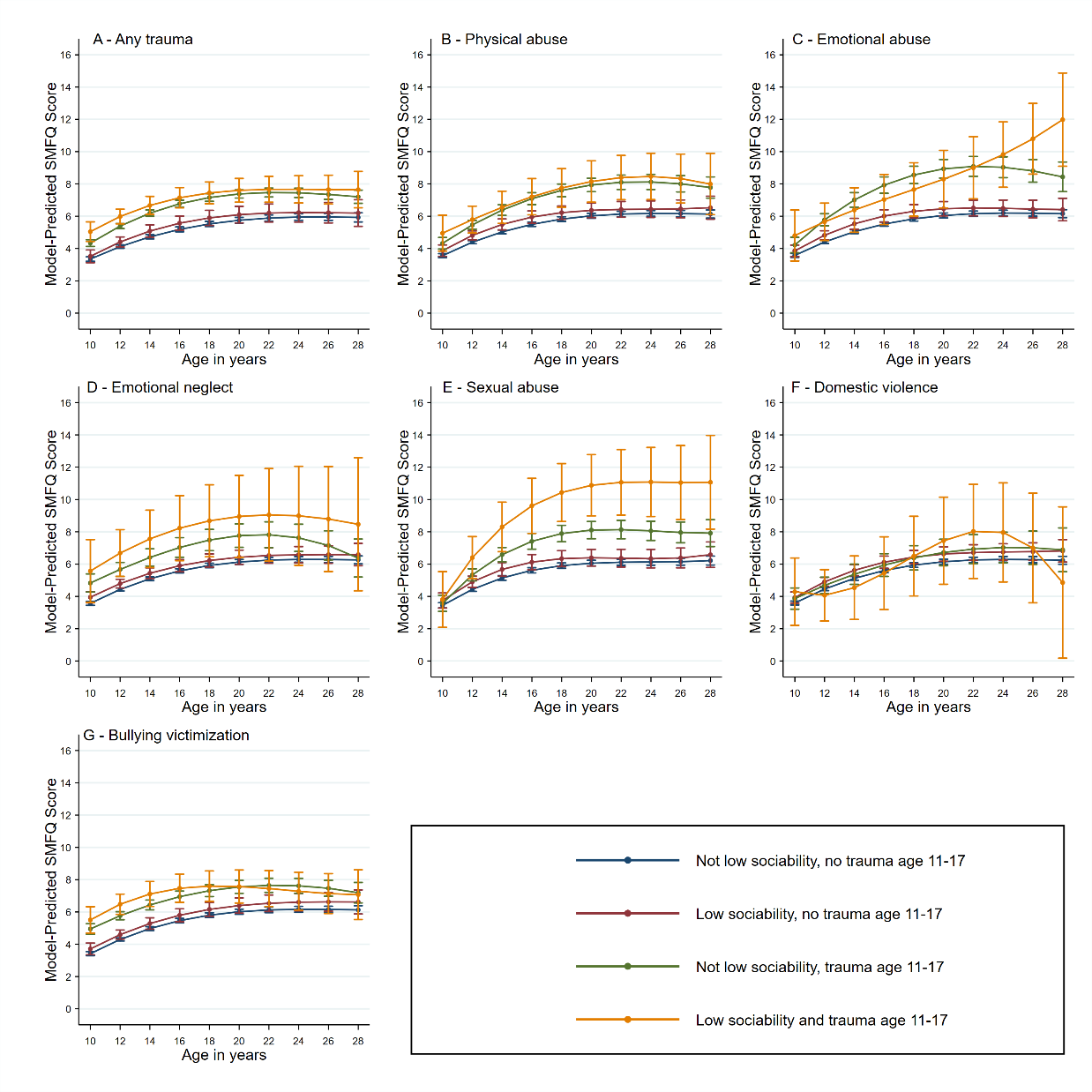

#### 2.7 Supplementary Results Figure S7: Trajectories of depressive symptoms between age 10 and 28 according to being in the top decile of the autism PGS and each trauma measure.

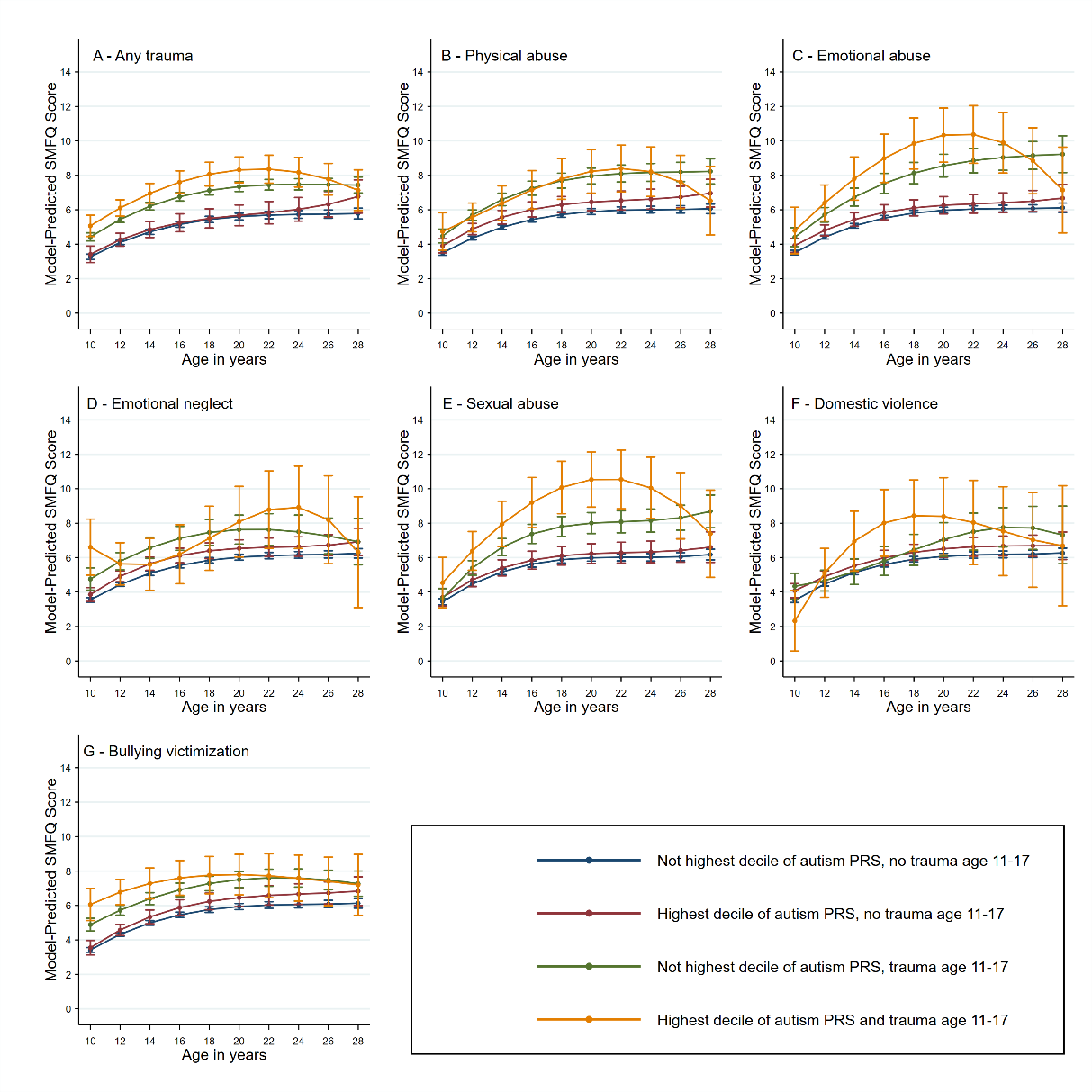

#### 2.8 Supplementary Results Figure S8: Odds ratios across autism polygenic risk p value thresholds for autism diagnoses and autism associated trait measures.

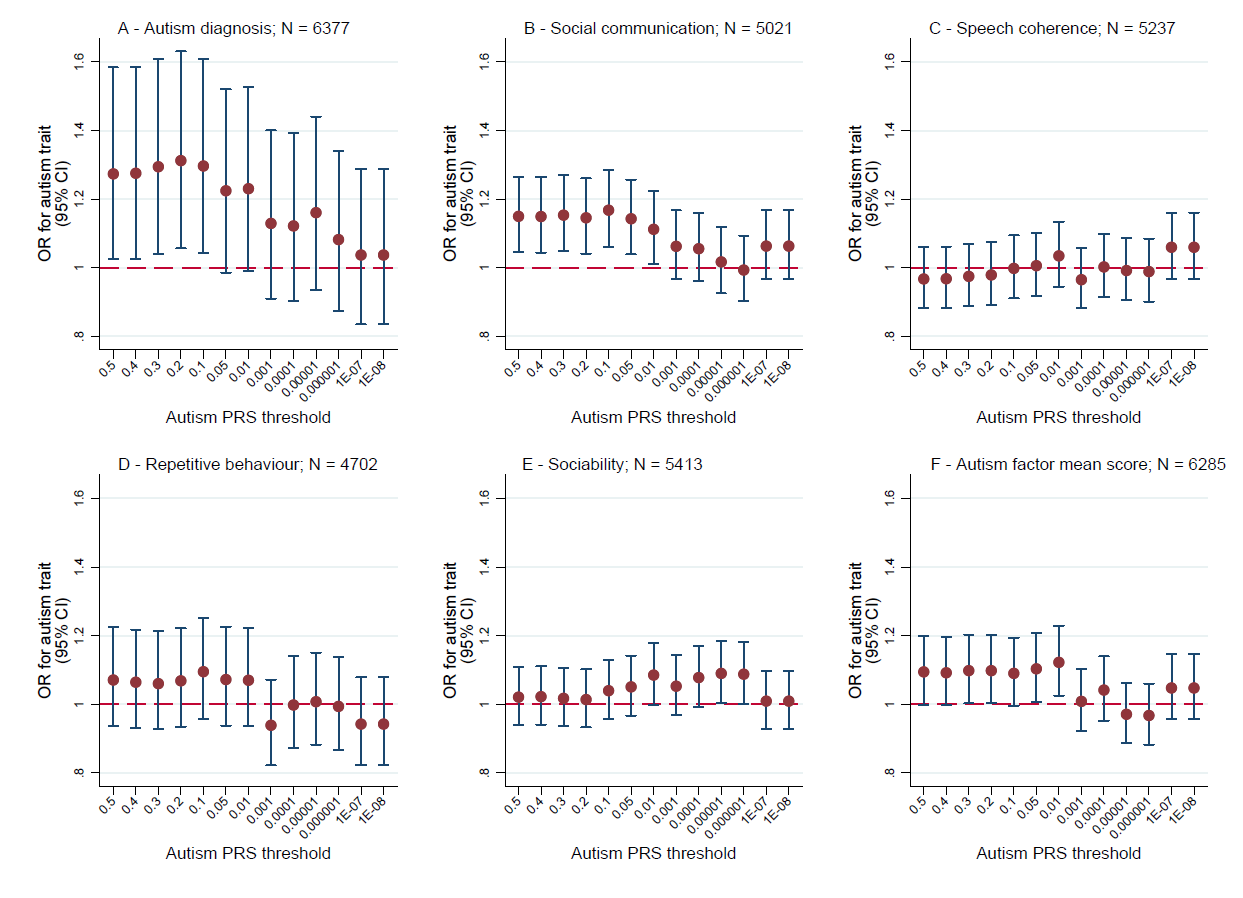
